# Stepwise Posterior-Based Arthroscopic Release for Severe Elbow Stiffness: Intraoperative Identification of a Critical Posteromedial Restraint

**DOI:** 10.64898/2026.02.06.26345629

**Authors:** Shinsuke Sakoda, Manabu Yamashita, Hiroto Kumagae, Arata Yoshida, Kimiaki Kawano

**Author notes:** **Corresponding Author** Shinsuke Sakoda, MD, Department of Sports Medicine, Ashiya Central Hospital, 283-7 Yamaga, Ashiya-machi, Onga-gun, Fukuoka 807-0141, Japan. **Funding Statement** No external funding was received for this study. **Ethical Approval** This retrospective multicenter study was approved by the institutional review board of Ashiya Central Hospital, which served as the central institutional review board (Approval No. ACH-12-2). **Patient Consent** The requirement for informed consent was waived by the institutional review board due to the retrospective nature of the study.

## Abstract

**Background:** Arthroscopic release for elbow stiffness is considered a minimally invasive and effective treatment. However, the extent to which each intraoperative step contributes to improvement in range of motion (ROM) has not been well investigated.

**Purpose:** To sequentially evaluate the relationship between intraoperative surgical steps and changes in elbow ROM during arthroscopic release for severe elbow stiffness, and to identify the key procedural stage contributing most significantly to ROM improvement.

**Methods:** Five elbows in five patients with severe elbow stiffness following fracture or dislocation were retrospectively reviewed. Arthroscopic release was performed using a stepwise posterior-based approach, starting from the posterior soft-spot portal, followed by exposure of the olecranon fossa and progression into the posteromedial compartment. Changes in elbow ROM were assessed at each intraoperative step, and ROM at final follow-up was also evaluated.

**Results:** All patients demonstrated improvement in elbow ROM at final follow-up. Intraoperative ROM improvement did not occur in a continuous manner but rather in a stepwise fashion. Gradual improvement was observed with establishment of the posterior and posteromedial working spaces, followed by the most substantial increase in ROM immediately after release of the soft tissue attached to the posterior aspect of the humeral medial epicondyle. Although the maximum ROM achieved intraoperatively was not fully maintained at final follow-up, no patient experienced deterioration to preoperative ROM levels.

**Conclusions:** In arthroscopic release for severe elbow stiffness, improvement in elbow ROM occurs in a stepwise rather than continuous pattern. Release of the posteromedial structures attached to the posterior aspect of the humeral medial epicondyle may represent a critical turning point contributing significantly to ROM improvement.

## Introduction

Elbow stiffness is a common condition following trauma or surgical intervention and can lead to substantial limitations in activities of daily living. ^9^ The functional range of motion required for the elbow joint is generally considered to be from 30° of extension to 130° of flexion, and restriction beyond this range represents a clinically significant problem. ^6^

Open surgical release has traditionally been performed for the treatment of elbow stiffness. ^6, 8, 9^ In recent years, however, arthroscopic elbow release has been increasingly reported as an effective alternative that achieves meaningful improvement in range of motion while minimizing surgical invasiveness. ^1, 4, 5, 10, 11^

Despite these advantages, elbow arthroscopy remains technically demanding because of the close proximity of neurovascular structures. ^3, 7^ In cases of severe stiffness, initial access to the anterior compartment itself may be particularly challenging. Consequently, standardized guidelines regarding which structures should be released and in what sequence have not been clearly established. Moreover, the relative contribution of each intraoperative step to the improvement in elbow range of motion has not been well elucidated. ^1, 10^

In patients with severe elbow stiffness, not only the anterior capsule and intra-articular scar tissue but also posterior and posteromedial structures may contribute to restriction of motion. In particular, the posteromedial structures attached to the posterior aspect of the humeral medial epicondyle are anatomically positioned to limit elbow extension, ^2, 4, 5^ yet their pathological significance has not been sufficiently investigated.

The purpose of this study was to clarify, through continuous intraoperative assessment of elbow range of motion during arthroscopic release for severe elbow stiffness, at which specific surgical stages meaningful gains in motion are achieved. Furthermore, we aimed to evaluate the relationship between release of the posteromedial structures attached to the posterior aspect of the humeral medial epicondyle and subsequent improvement in range of motion, and to discuss the role of posterior restraint mechanisms in severe elbow stiffness.

## Materials and Methods

### Patients

This study included five patients (five elbows) with severe elbow stiffness who underwent arthroscopic release performed by the senior author at two institutions. In all cases, elbow stiffness developed after surgical treatment for an elbow fracture or dislocation and failed to improve despite sufficient conservative management (Table 1).

**Table 1.** Patient demographics and initial injury characteristics.

|  | **Age (years)** | **Sex** | **Initial injury** |
| --- | --- | --- | --- |
| Case 1 | 11-15 | Female | Monteggia fracture |
| Case 2 | 46-50 | Male | Capitellar fracture |
| Case 3 | 41-45 | Male | Open fracture around the elbow |
| Case 4 | 11-15 | Female | Distal humeral condylar fracture |
| Case 5 | 11-15 | Male | Fracture-dislocation of the elbow |

### Stepwise Arthroscopic Release Technique

In this procedure, a stepwise posterior-based approach was adopted, anticipating difficulty with initial anterior access in severely stiff elbows. Even in cases of severe stiffness, entry into the joint can be achieved relatively safely from the posterior soft spot toward the posterior radiocapitellar joint while avoiding neurovascular structures.

#### 1. Posterior Approach

Patients were placed in either the lateral decubitus or prone position with the elbow maintained at approximately 90° of flexion. The procedure was initiated through the posterior soft spot portal, approaching the posterior radiocapitellar joint. Because proliferative inflammatory synovium and scar tissue frequently limited visualization, osseous landmarks were used as reference points. Scar tissue was sequentially debrided from the lateral aspect of the olecranon toward the olecranon fossa to create a working space. At this stage, little to no obvious improvement in elbow range of motion was typically observed.

#### 2. Release of the Posterior Capsule

The working space within the olecranon fossa was gradually expanded, and the posterior capsule was released from the humerus as extensively as possible until exposure of the muscle belly was achieved. This step often resulted in gradual improvement in flexion– extension motion; however, sufficient restoration of range of motion was not achieved at this stage alone.

#### 3. Expansion Toward the Posteromedial Compartment

Subsequently, the working space was extended in the posteromedial direction using the medial aspect of the olecranon and the posterior medial humerus as landmarks. Dissection was advanced toward the posterior aspect of the humeral medial epicondyle. To minimize the risk to the ulnar nerve, use of a shaver and radiofrequency device was limited, and stepwise dissection was primarily performed using a blunt rasp while carefully identifying bony landmarks.

#### 4. Exposure and Release of the Posteromedial Structures

After confirming the posterior surface of the humeral medial epicondyle, blunt dissection was performed along the bony surface of this area (Figure 1). Use of a shaver and radiofrequency device was restricted to minimal maneuvers required for visualization. Changes in elbow flexion–extension range of motion were assessed immediately after completion of this step.

**Figure 1.**
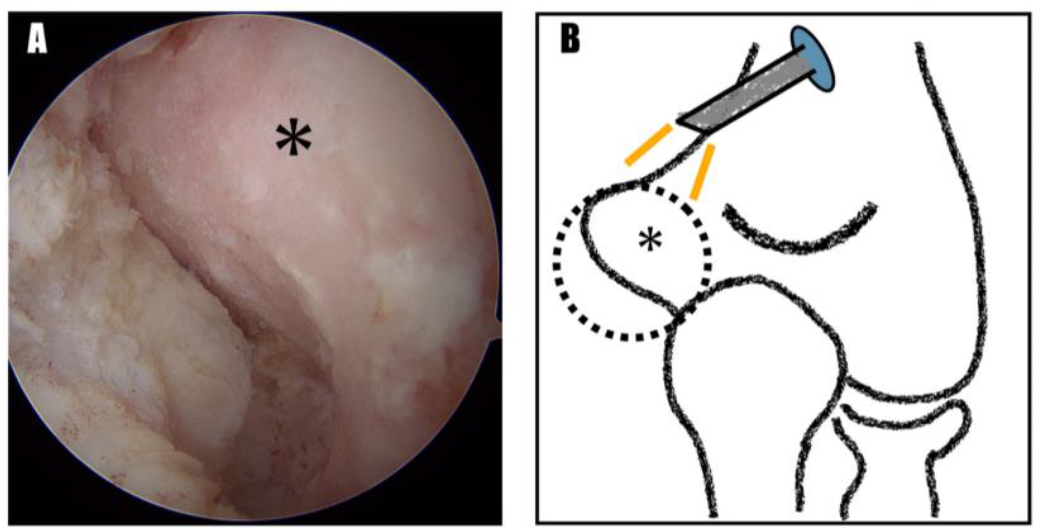
Arthroscopic view from a trans-triceps portal and schematic illustration of the posterior aspect of the medial epicondyle. (A) Arthroscopic image demonstrating the posterior surface of the humeral medial epicondyle (*). (B) Schematic illustration showing the arthroscopic approach to the posterior aspect of the medial epicondyle. The dotted circle indicates the target area corresponding to the arthroscopic field shown in panel A.

#### 5. Arthroscopic Visualization of the Anterior Compartment From Posterior

After completing posterior procedures, the arthroscope was advanced from the posterior compartment into the anterior compartment through the interval between the radiocapitellar joint and the ulna. Visualization from posterior to anterior confirmed that sufficient space had been created within the anterior joint compartment

#### 6. Creation of Anterior Portals Using the Outside-In Technique and Final Release

Once adequate space within the anterior compartment was confirmed, an anteromedial portal was created using the outside-in technique under posterior arthroscopic visualization. The anterior compartment was further expanded using a radiofrequency device and shaver. After sufficient working space was obtained, the arthroscope was switched to the anteromedial portal, and an anterolateral portal was established. Finally, the anterior capsule and surrounding scar tissue were released, and elbow range of motion was assessed.

### Postoperative Rehabilitation

Postoperatively, no external immobilization was used in any patient. Active and passive range-of-motion exercises were initiated as early as possible. Rehabilitation was continued with adjunctive treatments as needed according to pain and swelling, with the goal of maximizing postoperative improvement in elbow motion.

## Results

Preoperatively, elbow range of motion was markedly restricted in all patients. The mean duration of follow-up was 15.4 months (range, 3–26 months). At final follow-up, improvement in range of motion was observed in every case. Mean extension deficit improved from −35° preoperatively to −7° at final follow-up, and the arc of motion increased from 50° preoperatively to 108° at final follow-up (Table 2).

**Table 2.** Changes in elbow range of motion before surgery and at final follow-up.

|  | **Preoperative** | **Final follow-up** |
| --- | --- | --- |
| Extension deficit, mean (range) | -35 (-5 to -45) | -11 (-20 to 0) |
| Arc of motion, mean (range) | 50 (20 to 95) | 108 (85 to 120) |

Intraoperative improvement in range of motion did not occur in a continuous manner but rather in a stepwise fashion. Although gradual improvement was observed with the establishment of posterior and posteromedial working spaces, the greatest gain in range of motion was consistently achieved immediately after release of the soft tissues attached to the posterior aspect of the humeral medial epicondyle.

Although the maximum range of motion achieved intraoperatively was not fully maintained at final follow-up, no deterioration to preoperative levels or clinically significant recurrent stiffness was observed. No major complications, including neurologic injury or infection, occurred during the study period.

## Discussion

The most important finding of this study was that improvement in range of motion in severe elbow stiffness did not occur in a continuous manner but rather in a stepwise fashion, with a distinct turning point at a specific surgical stage. Although gradual gains in motion were observed during stepwise creation of posterior and posteromedial working spaces, the greatest and qualitatively different improvement in range of motion was consistently achieved immediately after release of the soft tissues attached to the posterior aspect of the humeral medial epicondyle.

In conventional arthroscopic elbow release procedures, surgical attention has primarily focused on release of the anterior or posterior capsule. ^1, 4, 5, 10, 11^ Detailed descriptions of surgical strategies extending into the posteromedial region remain limited. However, this does not necessarily reflect a lack of importance of the posteromedial structures; rather, it may be attributable to the absence of clearly described and widely shared techniques that enable safe access to this region in cases of severe stiffness. ^3, 7^

In patients with severe elbow stiffness, the anterior compartment is often functionally obliterated by dense scar tissue, rendering initial anterior access not merely technically demanding but, in some cases, fundamentally unfeasible. ^6, 8, 9^ Accordingly, the present study adopted a strategy that did not presuppose anterior entry but instead emphasized stepwise acquisition of intra-articular space from the posterior aspect.

Even in severely stiff elbows, although sufficient joint space is not initially present posteriorly, access to the joint could be achieved relatively safely by entering through the posterior soft spot toward the posterior radiocapitellar joint while avoiding major neurovascular structures. ^3, 7^ This surgical strategy was not originally designed to target a specific ligamentous structure; rather, it evolved pragmatically from initiating the procedure at the safest possible entry point and progressively expanding the working space from posterior to medial.

Notably, completion of posterior release up to the olecranon fossa resulted in only limited improvement in range of motion and was insufficient for effective contracture release. ^12^ Recognition that the posteromedial structures remained untreated at this stage prompted further medial expansion. Immediately after blunt release of the soft tissues attached to the posterior aspect of the humeral medial epicondyle along the bony surface, range of motion improved markedly and in a manner distinct from prior stages.

This intraoperative finding suggests that the posteromedial capsuloligamentous complex adjacent to the posterior aspect of the medial epicondyle may function as a final and dominant mechanical restraint in severe elbow stiffness. These structures have been described in anatomical and biomechanical studies as posteromedial components that include the posterior oblique ligament (POL). ^2, 4, 5^ The POL has been reported to contribute to restraint in terminal elbow extension. ^2, 5^ The marked improvement in extension observed in the present study is consistent with these biomechanical insights and is most reasonably interpreted as functional release of the posteromedial capsuloligamentous complex, including structures corresponding to the posterior oblique ligament (POL). It should be noted, however, that this structure was not visualized intraoperatively as a discrete ligament, and identification of the POL in this study is based on functional interpretation inferred from the anatomical attachment site and the corresponding changes in range of motion.

The surgical logic demonstrated in this study aligns with the stepwise approach commonly applied in arthroscopic procedures of other joints, in which safe working spaces are sequentially established to ultimately address the principal restraint structures.^1^

### Limitations

This study is limited by its small sample size and retrospective design. Nevertheless, the consistent intraoperative pattern of range-of-motion improvement observed across all cases supports the validity of the proposed surgical strategy.

### Conclusion

In arthroscopic release for severe elbow stiffness, a strategy that begins posteriorly and progresses in a stepwise manner appears to be rational and effective, rather than initiating the procedure from the anterior compartment. Functional release of the posteromedial restraint, corresponding to the POL complex, is associated with substantial improvement in range of motion, while subsequent anterior release serves as a finishing step. This technique may represent a reproducible surgical strategy grounded in the fundamental principles of stepwise arthroscopic intervention.

## Data Availability

All data produced in the present study are available upon reasonable request to the authors

## Notes

**Conflict of Interest Statement** The authors declare no conflicts of interest related to this study.

### Competing Interest Statement

The authors have declared no competing interest.

### Funding Statement

None.

### Author Declarations

Ethics Committee of Ashiya Central Hospital gave ethical approval for this work.

